# An Analysis of Patterns of Distribution of Buprenorphine in the United States using ARCOS, Medicaid, and Medicare Databases

**DOI:** 10.1101/2022.08.22.22279092

**Authors:** Zhi-Shan Hsu, Justina A. Warnick, Tice R. Harkins, Briana E. Sylvester, Neal K. Bharati, leTausjua B. Eley, Stephanie D. Nichols, Kenneth McCall, Brian J. Piper

**Author notes:** **Corresponding Author Information** Justina A. Warnick, 617 Highland Ave, Lewistown PA 17044. **Prior Presentations**: The findings from this manuscript have been previously presented in poster format at the Geisinger Commonwealth School of Medicine Spring Student Research Symposium in Scranton, PA in April 2021. A preprint was submitted to MedRxiv on 8/22/2022.

## Abstract

**Purpose:** Opioid overdose remains a problem in the United States despite pharmacotherapies, such as buprenorphine, in the treatment of opioid use disorder (OUD). This study characterized changes in buprenorphine use.

**Methods:** Using the Drug Enforcement Administration’s ARCOS, Medicaid and Medicare claims databases, patterns in buprenorphine usage in the United States from 2018 to 2020 were analyzed by examining percentage changes in total grams distributed and changes in grams per 100K people in year-to-year usage based on zip-code and state levels.

**Results:** For ARCOS from 2018-19 and 2019-20, total buprenorphine distribution in grams increased 16.2% and 12.6% respectively. South Dakota showed the largest state-wide percentage increase in both 2018-19 (66.1%) and 2019-20 (36.7%). From 2018-19, the zip codes ND-577 (156.4%) and VA-222 (−82.1%) had the largest and smallest percentage changes respectively. From 2019-20, CA-932 (250.2%) and IL-603 (−36.8%) were the largest and smallest respectively. In both 2018-19 and 2019-20, PA-191 had the second highest increase in grams per 100K while OH-452 was the only zip code to remain in the top 3 largest decreases in grams per 100K in both periods. Among Medicaid patients in 2018, there was a 1,932.1-fold difference in prescriptions per 100k Medicaid enrollees between Kentucky (12,075.5) and Nebraska (6.25). Among Medicare enrollees in 2018, family medicine physicians and other primary care providers were the top buprenorphine prescribers.

**Conclusions:** This study identified overall increases in buprenorphine availability but also pronounced state-level differences. Such geographic analysis can be used to discern which public policies and regional factors impact buprenorphine access.

**Key Points:**

- Total buprenorphine distribution in grams increased 16.2% and 12.6% for 2018-2019 and 2019-2020 ARCOS data, respectively
- On a state level, from 2020-21, the majority of states were decreasing buprenorphine distribution as reported by ARCOS
- The state with the highest buprenorphine usage among Medicaid patients in both 2018 and 2019 was Kentucky
- Family medicine physicians were the top buprenorphine prescribers to Medicare enrollees in 30 states
- Maine had the largest number of buprenorphine prescriptions to Medicare enrollees

**Plain Language Summary:** Opioid overdose remains a problem in the United States despite medical therapy, such as buprenorphine, in the treatment of opioid use disorder (OUD). This study aimed to describe changes in buprenorphine use. Using the Drug Enforcement Administration’s ARCOS, Medicare, and Medicaid databases, patterns in buprenorphine usage in the United States from 2018 to 2020 were analyzed based on zip-code and state levels. For ARCOS from 2018-19 and 2019-20, total buprenorphine distribution in grams increased 16.2% and 12.6% respectively.South Dakota showed the largest state-wide percentage increase in both 2018-19 (66.1%) and 2019-20 (36.7%). From 2018-19, the zip codes ND-577 (156.4%) and VA-222 (−82.1%) had the largest and smallest percentage changes respectively. From 2019-20, CA-932 (250.2%) and IL-603 (−36.8%) were the largest and smallest respectively. Among Medicaid patients in 2018, there was a 1,932.1-fold difference in prescriptions per 100k Medicaid enrollees between Kentucky (12,075.5) and Nebraska (6.25). Among Medicare enrollees in 2018, family medicine physicians and other primary care providers were the top buprenorphine prescribers. This study found overall increases in buprenorphine availability but also pronounced state-level differences. Such geographic analysis can be used to determine which public policies and regional factors impact access to buprenorphine.

**Ethics Statement:** Hereby, I Justina A. Warnick consciously assure that for the manuscript “An Analysis of Patterns of Distribution of Buprenorphine in the United States using ARCOS, Medicaid, and Medicare Databases” the following is fulfilled:

1. This material is the authors’ own original work, which has not been previously published elsewhere aside from preprint server MedRxiv.
2. The paper is not currently being considered for publication elsewhere.
3. The paper reflects the authors’ own research and analysis in a truthful and complete manner.
4. The paper properly credits the meaningful contributions of co-authors and co-researchers.
5. The results are appropriately placed in the context of prior and existing research.
6. All sources used are properly disclosed (correct citation). Literally copying of text must be indicated as such by using quotation marks and giving proper reference.
7. All authors have been personally and actively involved in substantial work leading to the paper and will take public responsibility for its content.

I acknowledge that the violation of the Ethical Statement rules may result in severe consequences.

**Graphical Abstract:** 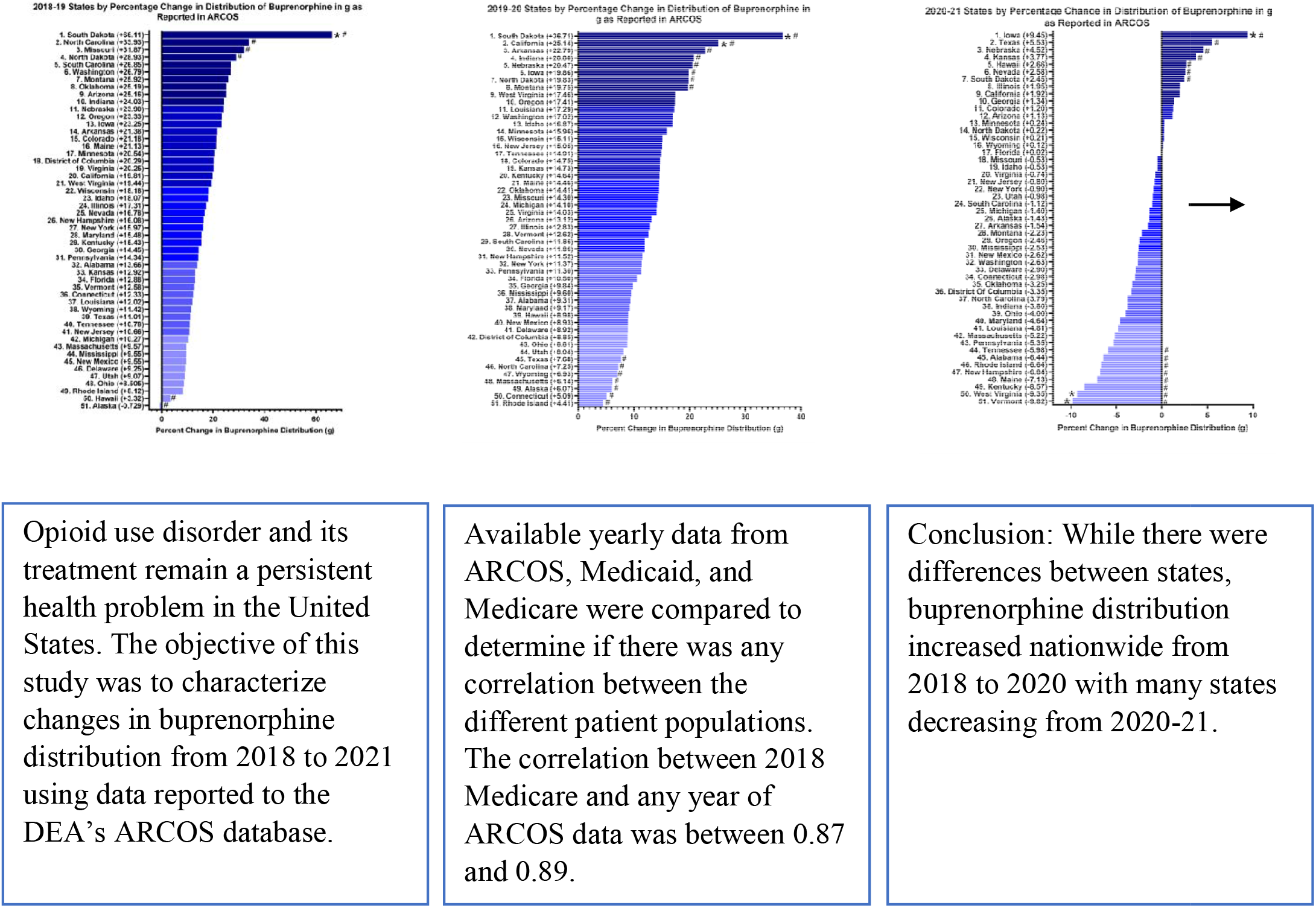

## Introduction

Drug overdoses during the largely iatrogenic opioid epidemic have claimed the lives of over one million Americans^1^. Of the 70,630 drug related overdose deaths reported to the Center for Disease and Control (CDC) in 2019, 72.9% were the result of opioid use. In addition to the appreciable mortality rate, the overall economic cost (healthcare, criminal justice, and workplace costs) of the opioid crisis has risen from $11.8 billion in 2001 to $78.5 billion in 2016^2,3^. In response to the opioid epidemic, the Centers for Disease Control and Prevention (CDC) is focused primarily on prevention. The CDC currently aims to monitor trends in opioid use, advance research to identify which areas of the country need support and improve opioid prescribing and safety through provider education. While prevention of opioid use disorder (OUD) is necessary to combat the epidemic, it is equally important to treat the estimated two million Americans currently living with OUD^4^.

The current gold standard for treatment of OUD is opioid agonist medications (e.g., buprenorphine and methadone)^5^. Treatment of OUD with buprenorphine has been demonstrated to reduce the risk of opioid drug related death by as much as 50%^6^. Buprenorphine is a mu receptor partial agonist, kappa receptor agonist, and delta receptor antagonist, which mitigates symptoms of opioid dependency and prevents withdrawal symptoms among individuals with OUD^7,8^. As a partial agonist of opioid receptors, buprenorphine exerts weaker opioid effects than that of other full agonist opioids such as morphine, and consequently has less tendency for overdose. Buprenorphine’s use in the treatment of OUD has predicated on the fact that partial agonist activity is safer than methadone, a full agonist opioid dependence therapy whose availability for OUD is limited to specialized facilities. When prescribed at fixed, high dosages (greater than 7 mg daily), buprenorphine proves an effective therapy for decreasing illicit opioid use^5^.

Even though the efficacy of treatment has been well-established, the implementation of prescribing buprenorphine has not been without setbacks and obstacles. In the past two years, however, policy changes have been moving in the direction of increasing access. The Coronavirus (COVID-19) pandemic has accelerated the expansion of telehealth services through modifications to payment, privacy, and licensing regulations^9^. Among the changes in licensing regulations was the decision to waive the Ryan Haight Act’s mandate that an in-person examination be performed prior to the prescription of buprenorphine^10^. The removal of the Ryan Haight Act’s in-person examination mandate allowed for the initial consultation of a patient with OUD to be conducted via telemedicine, thereby significantly reducing a barrier to access that affected patients with OUD who lacked means of transportation or a licensed provider in their county. Although this was a significant improvement, there remain serious gaps and inequities in treatment access and availability based on geographic location and race^11^. For example, the prevalence of OUD is similar among Black and White adults, but Black patients are less likely to be prescribed buprenorphine at an outpatient visit^12^. This may be partially due to the availability of prescribing providers. Currently, advanced practice pharmacists with prescribing authority struggle with policy constraints associated with buprenorphine use. There have been easement of restraints including removal of the lengthy educational requirements to obtain the X-waiver which allows a provider to prescribe buprenorphine. With less training requirements, more providers are likely to apply, but nonetheless the X-waiver itself remains in place. Furthermore, clinical pharmacists remain unable to prescribe buprenorphine as they are unable to obtain an X-waiver – presenting a major missed opportunity to effectively increase access^13^.

With the COVID-19 pandemic impacting many aspects of healthcare in 2020, there is reason to ask how buprenorphine usage may have been affected. The increased use of telemedicine during the pandemic along with emergency easing of the Ryan Haight Act and DEA requirements to obtain a separate license in each state in which they practice enhanced access to treatment for OUD^14^. Specifically, use of buprenorphine has substantially increased in recent years, partially due to Medicaid expansion under the Affordable Care Act^11^. The objective of this study was to expand upon existing knowledge regarding geographic disparities in the distribution and access to buprenorphine as well as consider coinciding modifications to Medicaid, Medicare, and telemedicine regulations.

## Methods

### Procedures

Information on annual distribution of buprenorphine was extracted from the Drug Enforcement Administration’s Automation of Reports and Consolidated Orders System (ARCOS). Retail Drug Summary Reports on the ARCOS^15^ reports by year and by state for a given controlled substance. Using the 2018-20 reporting periods, all data listed under “buprenorphine” were used. This comprehensive database includes private and public (e.g., Veterans’ Affairs) pharmacies and has been used in prior research^16^. Data was also obtained from the State Drug Utilization Data (SDUD) database on the Medicaid website^17^. Using the 2018 and 2019 reporting periods, all data listed under “buprenorphine” were used. The National Drug Codes (NDCs) used were listed in Appendix A. Using the 2018 reporting period, the most recent available when analyses were completed, all data listed under “buprenorphine” were obtained ^18^. Mono- and combo products were both included and not separated in this analysis.Data on Medicaid and Medicare was extracted from data.cms.gov. The number of buprenorphine prescriptions to patients on Medicare was extracted from the years 2018 and 2019.Buprenorphine prescriptions to patients on Medicare were obtained for 2018 which was the most recent available. This research was approved by the Geisinger IRB as exempt.

### Data Analysis

For each state, ARCOS categorizes drug usage by zip code tabulation areas (ZCTAs), which are 3-digit zip code prefixes. For example, the grams of buprenorphine distributed in zip code 18510 would be merged with the distribution of all other zip codes beginning with 185 and reported as “185.” To create geographic heat maps for this data, the open-source software QGIS was used. While ARCOS contained distribution data for Washington D.C., Puerto Rico, and other United States territories^19^, they were excluded from detailed analysis. The periods assessed were 2018-19 and 2019-20. The percentage changes in buprenorphine distribution were then calculated using the formulas:

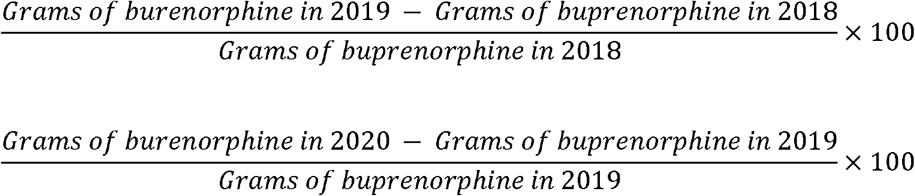

for 2018-19 and 2019-20 respectively. Because percentage change can artificially inflate the magnitude of changes when the initial value is small, zip codes with initial buprenorphine distributions less than 500 g were excluded from analysis of percentage change. Absolute changes in buprenorphine distribution were calculated by finding the difference in grams of buprenorphine per 100K population reported from 2018-19 and 2019-20. They will be reported in units of mg/100 people. Individual 3-digit zip code regions will be referred to by their 2-letter state abbreviation followed by the ZCTA (e.g., VA-222 refers to all zip codes beginning with 222, which ARCOS reports is in Virginia). To calculate 95% confidence intervals, standard deviation (SD) was calculated using the *STDEV*.*P* function in Excel and then margin of error (MoE) calculated using with an ^α^ = 0.05 using this equation:

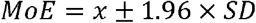

The 95% CI was calculated as one MoE above and below the mean of a given set of percentage or absolute changes in buprenorphine distribution.

Medicaid data was normalized for prescriptions per 100,000 Medicaid enrollees by state using the enrollment data on the Medicaid website^20^. The percentage and absolute changes in buprenorphine prescriptions were then calculated the same way as the ARCOS percentage changes. The data was then plotted using GraphPad Prism and statewide geographic maps using the Geographic Heat Map application for Excel. Standard deviation and confidence intervals for absolute change were calculated using the same method as for ARCOS data.

The Medicare data was plotted using Microsoft Excel to show each state’s annual total buprenorphine prescription for 2018. The data was then normalized for prescriptions per 100,000 Medicare enrollees by state using the enrollment data on the Kaiser Family Foundation website^21^. The data was then sorted by provider type to determine the most common providers to prescribe Buprenorphine by state and overall. The data was then plotted using GraphPad Prism.

## Results

### ARCOS

On a national level, there was an increase in buprenorphine distribution. From 2018-19, there was a +16.2% increase in total grams of buprenorphine distributed and an increase in 196 mg/100 people. From 2019-20, there were increases of 12.6% and 178 mg/100 people. After calculating the percentage change in buprenorphine usage by zip code from 2018 to 2019 (Figure 1a, Figure 1b), it was noted that the ND-577 (156.4%, an increase from 598.8 g in 2018 to 1,408.7 g in 2019) and the VA-222 (−82.1%, a decrease from 4,187.3 g in 2018 to 748.3 g in 2019) had the largest and smallest percentage changes respectively. In comparison, from 2019-10 (Figure 1c), the CA-932 (250.2%, an increase from 2,296.3 g in 2019 to 8,042.1 g in 2020) and Illinois zip code 603 (−36.8%, a decrease from 1,457.6 g in 2019 to 920.6 g in 2020) were the largest and smallest respectively. From 2019-20, there is a cluster of zip codes in southern and eastern California with some of the largest percentage increases in buprenorphine distribution. This same cluster is not nearly as apparent from 2018-19. From 2020-21, though, there was a nationwide trend toward decreasing distribution of buprenorphine (Figure 1d). In both 2018-19 and 2019-20, on a state level (Figure 2a, 2b, Supplemental Figure 1a, 1b), South Dakota had a percentage increase that fell outside of a 95% CI from the mean. However, based on the geographic analysis, many of the South Dakota zip codes in both time periods had a buprenorphine distribution less than 500 g. Maine, Kentucky, and West Virginia had absolute increases outside of a 95% CI from the mean in both time intervals. Given the geographic pattern observed in 2019-20 on the zip code level, it is notable that the state-level percentage increase in California fell outside of a 95% CI from the mean. On a state level, from 2020-21 (Figure 2c, 2d), the majority of states were decreasing buprenorphine distribution. West Virginia continued its pattern of increasing distribution in mg/100 people and Iowa continued to rank highly in percentage change in grams distributed.

**Figure 1.**
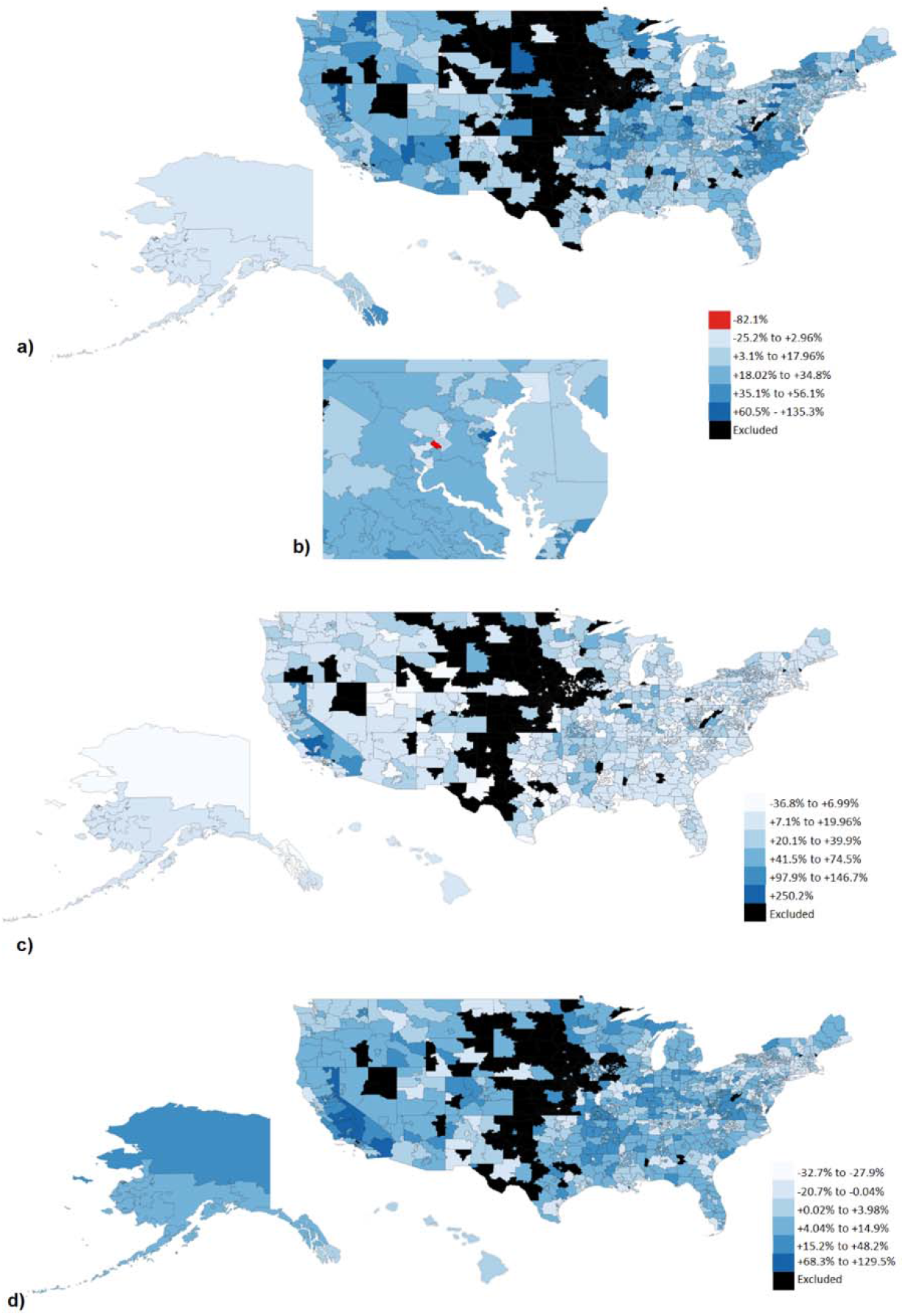
a) Percent change in buprenorphine by weight distributed between 2018 and 2019 by three-digit zip code from the Drug Enforcement Administration’s Automated Reports and Consolidated Orders System (ARCOS). Zip codes with a total distribution in 2018 of less than 500 g were excluded (black). b) The only zip code with a decreased value from 2018-19 (–82%) was VA-222, which is shown in an expanded view. c) Percent change in buprenorphine by weight distributed between 2019 and 2020. Zip codes with a total distribution in 2019 of less than 500 g were excluded (black). The only zip code with a value of +250% was CA-932. d) Percent change in buprenorphine distribution by weight between 2020 and 2021. Zip codes with a total distribution in 2020 of less than 500 g were excluded (black).

**Figure 2.**
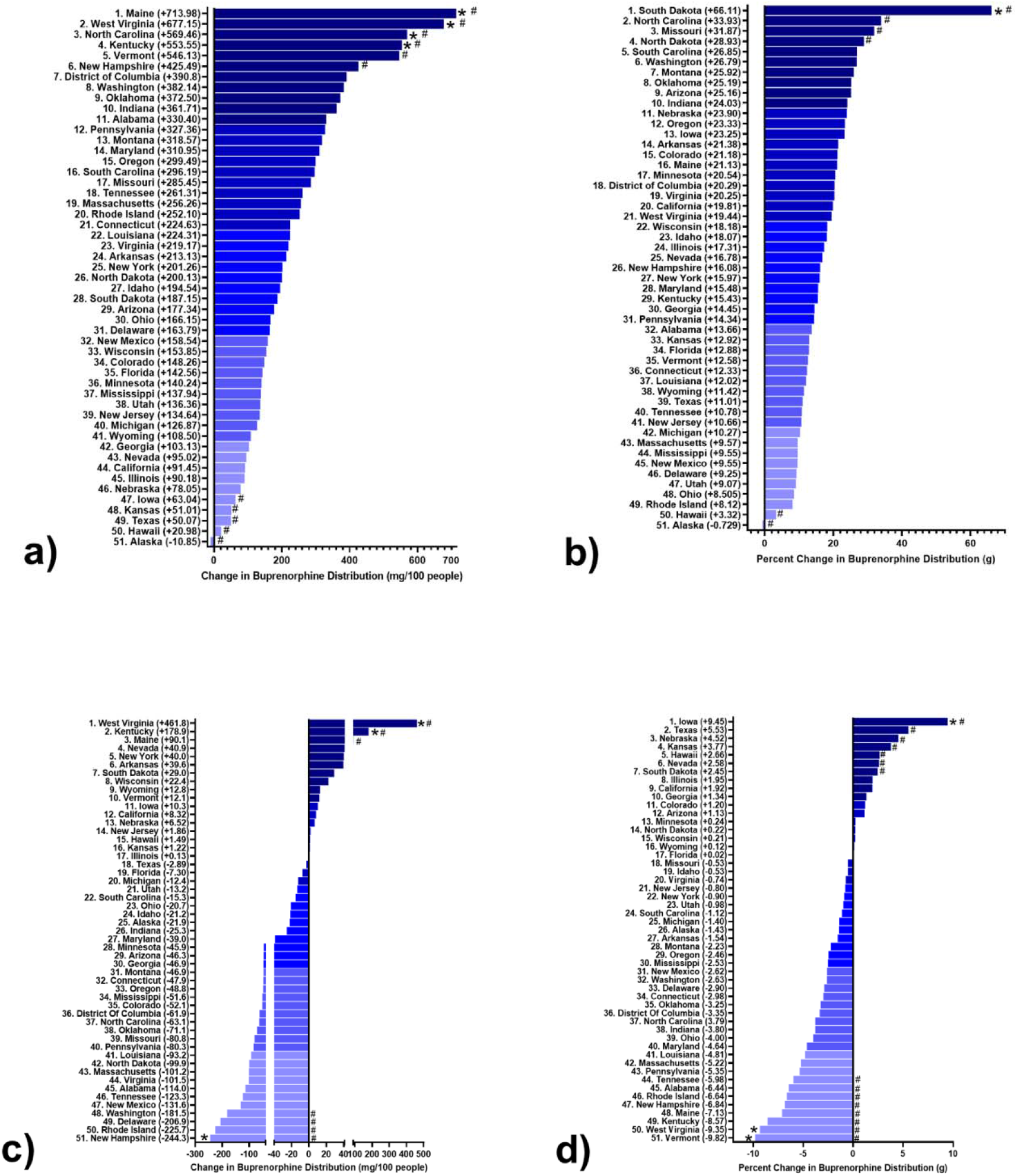
Change in state buprenorphine distribution in mg per 100 people from ARCOS database a) and percentage change in total grams of buprenorphine distributed per state b) from 2018-19. Percentage change was calculated using the total grams of buprenorphine distributed per state. States that fall outside of a 95% confidence interval from the mean percentage (17.8) and mean change in mg per 100 people (234.6) were noted with an asterisk (*). Change in state buprenorphine distribution in mg per 100 people c) and percentage change in total grams of buprenorphine distribution per state d) from ARCOS database for 2020-21. States that fall outside of a 95% confidence interval from the mean percentage (−1.73) and mean change in mg per 100 people (−31.2) were noted with an asterisk (*). A pound (#) sign indicates states that were greater than 1 SD from the mean.

### Medicaid

The state with the highest buprenorphine usage among Medicaid patients in both 2018 and 2019 was Kentucky (Supplemental Figure 2). In terms of change in usage from 2018-19 (Figure 3a, 3b), the largest absolute increase and decrease in buprenorphine prescriptions were Vermont and New Hampshire respectively, while the largest percentage increase and decrease were Michigan and tied between Wyoming and Nebraska respectively. Michigan only had the 17^th^ largest absolute increase in buprenorphine prescriptions while Vermont’s percentage change was the 4^th^ highest despite its large absolute change. New Hampshire’s percentage change was also ranked 40 out of 50, the 11^th^ largest percentage decrease. Wyoming and Nebraska also saw a very small number of buprenorphine prescriptions, so their large percentage changes appear to be inflated. When considering both absolute and percentage decreases, Louisiana ranked highly in both.

**Figure 3.**
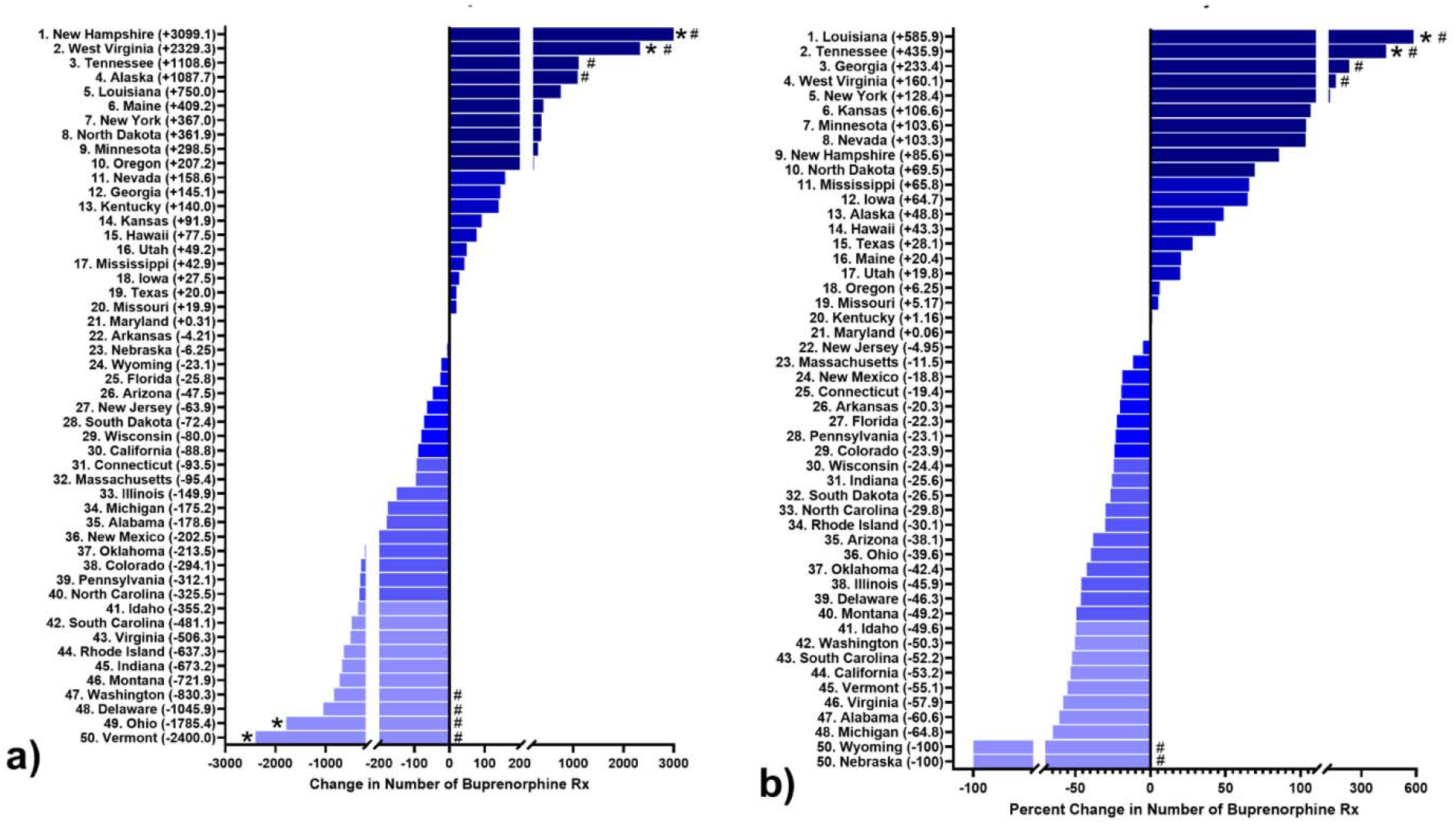
a) Absolute change in number of buprenorphine prescriptions per 100,000 Medicaid enrollees by state. States that fall outside of a 95% CI from the mean of the changes in prescriptions per 100K enrollees (21.9) are annotated with an asterisk (*). b) Percentage change in number of buprenorphine prescriptions per 100,000 Medicaid enrollees by state. States that fall outside of a 95% CI from the mean of percentage changes in number of prescriptions per 100K enrollees (17.1) are annotated with an asterisk (*) A pound (#) sign indicates states that were greater than 1 SD from the mean.

### Medicare

Using Medicare’s claims for buprenorphine in 2018, the goal was to find any potential patterns among providers and buprenorphine usage. Nationwide, family medicine physicians were the top prescribers of buprenorphine among Medicare patients (Supplemental Figure 3). When the providers were broken down to general providers and specialists (Supplemental Figure 4), there was a large drop from the top specialist, psychiatry, to the rest of the top five. When comparing the top prescribers in each state, family medicine physicians were the top prescribers in thirty states (Table 1). For all Medicare enrollees nationally, the state with the largest number of buprenorphine prescriptions was Maine (Supplemental Figure 5). Maine, Vermont, Massachusetts, and West Virginia all fell outside of a 95% CI from the mean.

**Table 1.** States categorized by top buprenorphine prescriber to Medicare enrollees in 2018. For each state, the specialty/provider type with the largest number of buprenorphine prescription claims in 2018 was found.

| Top Buprenorphine-Prescribing Specialty | States |
| --- | --- |
| Family Practice (30) | Alaska, Alabama, California, Colorado, Delaware, Florida, Iowa, Illinois, Indiana, Kansas, Kentucky, Louisiana, Maine, Michigan, Minnesota, Mississippi, New Hampshire, New Mexico, New York, Ohio, Oklahoma, Oregon, Pennsylvania, South Carolina, Tennessee, Texas, Utah, Vermont, Washington, West Virginia |
| Psychiatry (8) | Connecticut, Georgia, Hawaii, Missouri, North Carolina, Nevada, Wisconsin, Wyoming |
| Internal Medicine (5) | Massachusetts, Maryland, New jersey, Rhode Island, Virginia |
| Nurse Practitioner (4) | Arizona, Idaho, Montana, North Dakota |
| Anesthesiology (1) | Arkansas |
| Physician Assistant (1) | South Dakota |
| Pain Management (1) | Nebraska |

When examining all the datasets used in this analysis (Table 2), there was a strong and expected correlation between data from the same source across different years. Across datasets, there was a moderate correlation between Medicare and Medicaid. There was a larger correlation between 2018 Medicare with all four years of ARCOS data than either 2018 or 2019 Medicaid with the ARCOS data. With this in mind, it is notable that West Virginia had a large increase in absolute buprenorphine distribution from 2018-19 and 2019-20 in the ARCOS data and a large number of prescriptions in 2018 to Medicare enrollees.

**Table 2.** Correlation matrix of Medicare, Medicaid, and the Drug Enforcement Administration’s Automated Reports and Consolidated Orders System (ARCOS) data. Values above .328 were significant at *p* < .01.

|  | 2018<br>ARCOS<br>g/100K | 2019<br>ARCOS<br>g/100K | 2020<br>ARCOS<br>g/100K | 2021<br>ARCOS<br>g/100K | 2018<br>Medicaid<br>Rx/100K | 2019<br>Medicaid<br>Rx/100K | 2018<br>Medicare<br>Rx/100K |
| --- | --- | --- | --- | --- | --- | --- | --- |
| 2019<br>ARCOS<br>g/100K | 0.995 | 1 |  |  |  |  |  |
| 2020<br>ARCOS<br>g/100K | 0.989 | 0.997 | 1 |  |  |  |  |
| 2021<br>ARCOS<br>g/100K | 0.977 | 0.988 | 0.995 | 1 |  |  |  |
| 2018<br>Medicaid<br>Rx/100K | 0.588 | 0.591 | 0.599 | 0.605 | 1 |  |  |
| 2019<br>Medicaid<br>Rx/100K | 0.576 | 0.582 | 0.595 | 0.610 | 0.918 | 1 |  |
| 2018<br>Medicare<br>Rx/100K | 0.886 | 0.886 | 0.879 | 0.867 | 0.375 | 0.390 | 1 |

## Discussion

This report identified an overall increase in buprenorphine distribution and prescriptions but also sizable state-level disparities. These findings extend upon prior reports.^16^ Analysis of ARCOS usage data showed that in comparison to the changes from 2018-19, between 2019-20 there was an overarching pattern of increased buprenorphine usage across the entire United States. The highest percent increase in buprenorphine usage observed (over 500% increase in the 772 Alaska zip code region) was in the 2018-19 period. In 2019-20, the largest decrease (36.8% in the 603 Illinois zip code region) was more modest than the largest decrease seen in 2018-19 (100% in the Houston area). South Dakota had the largest percent increase in buprenorphine usage in both 2018-19 and 2019-20 on a state-wide basis.

ARCOS provides a comprehensive view of buprenorphine usage reported to the DEA among all patients. By using the buprenorphine usage data in 2018 and 2019 from Medicaid, the data from a subset of the ARCOS dataset was able to be further examined. This database provides a more granular perspective as it focuses specifically on Medicaid enrollees who meet certain requirements including low income, pregnancy, less than 18 years old, or disability.Kentucky had the highest buprenorphine usage in both 2018 and 2019. Kentucky has been one of the most successful states at implementing state insurance through the ACA. It has reduced the uninsured rate 58% from 2010-2019 and has seen a 161% increase in Medicaid enrollment since expanding eligibility. Furthermore, Kentucky has seen increases in preventative screening usage which may reflect an overarching theme of greater patient involvement in their own health, including seeking out medication assisted treatment with buprenorphine^22^. In contrast, Louisiana ranked highly when considering both absolute and percentage decreases. Like Kentucky, Louisiana also accepted federal Medicaid expansion and saw a comparable decrease (50%) in the uninsured rate but only a 71% increase in Medicaid enrollment. Many factors likely contribute to the differences in buprenorphine prescriptions between these two states^22,23^. One factor may be internet access, as more appointments and prescriptions are given via telehealth. According to BroadbandNow, Kentucky ranks 24th in the United States for internet coverage and availability compared to Louisiana which ranks 40^th^. An important caveat with the Medicaid data is that the buprenorphine product types were not divided (combination vs monotherapy). This may affect the data as some formulations are more commonly used for OUD versus pain management.

The Medicare database provided yet another lens through which to analyze the data as most Medicare enrollees are greater than 65 years of age or have end-stage kidney disease. When correcting for the number of enrollees, Maine has the largest amount of buprenorphine prescriptions. More notably however, is that, among specialists, the top providers all work in specialties where buprenorphine could be prescribed for the purpose of pain management rather than for OUD. Family medicine physicians were the top prescribers in thirty states including Maine. In contrast, addiction medicine physicians constituted only 2.94% of providers prescribing buprenorphine. This may reflect a difference in intended use (pain management vs OUD). Nevertheless, gathering information on buprenorphine prescription is important regardless of intended use as it can help understand patterns of use and diversion.

While the ARCOS, Medicaid, and Medicare databases provide complementary sources of information, caveats exist for each database. The data from Medicare and Medicaid shared similar limitations. Medicare data from 2019 and 2020 and Medicaid data from 2020 were not released at the time of this analysis. While patterns over three years could be identified for ARCOS, conclusions drawn from Medicare and Medicaid were more temporally limited. In addition, data from Medicare and Medicaid report for a limited, although important, subsets of the United States population: patients age 65+ and patients with low socioeconomic status that qualify for Medicaid coverage. However, within those groups, meaningful directions for further research and policy intervention can be identified. From the 2018-19 Medicaid data, Washington experienced the largest percent increase in buprenorphine usage while Wisconsin saw the largest percent decrease. Using these states as starting points, future studies could begin searching for reasons as to why these patterns exist. If, for example, buprenorphine is being under-prescribed due to policy/coverage changes or for external reasons such as local health trends, policy updates may provide meaningful change in treatment of OUD. From the 2018 Medicare data, it was noted that primary care providers were the most common prescribers in thirty states with the highest number of prescriptions being in Maine. As with Medicaid, this information could be used to direct evaluation of current Medicare policy toward buprenorphine.

One potentially large impact on the differences in buprenorphine usage from 2018-19 compared to 2019-20 could have been the COVID-19 pandemic and associated policies.Changes in buprenorphine usage could be compared to opioid-related deaths during the pandemic. If a region experienced an increase, factors such as financial instability and changes to transportation or access to pharmacies could play a role. If an area experienced a decrease in opioid-related deaths, those regions could potentially be used as models. Did those regions implement COVID-19 safety protocols differently in a way that maintained access to treatment centers or were there other factors that contributed to the decrease? Overall, a geographical analysis of buprenorphine usage can impact future policy and provide insight into the population level effectiveness of current practices in treating OUD. From this study, the fourteen states with a significant difference in the change in buprenorphine usage from 2018-19 and 2019-20 could be places to start detailed analysis of the potential effects of the pandemic. States with potential problems can be isolated and analyzed to ascertain the prescribing habits among specific provider levels and medical specialties and the opportunities for health or financial policy intervention.

## Data Availability

All data produced in the present study are available upon reasonable request to the authors

## Acknowledgements

The authors of this manuscript would like to acknowledge Iris Johnston and the Biomedical Research Club for their contributions. This research received no external funding.

---

**Supplemental Appendix A.** Buprenorphine National Drug Codes (NDC)

| **Proprietary Name** | **NDC Package Code** |
| --- | --- |
| Buprenorphine HCl | 0054-0176-13 |
| Buprenorphine HCl | 0054-0177-13 |
| buprenorphine hydrochloride and naloxone hydrochloride dihydrate | 0054-0188-13 |
| buprenorphine hydrochloride and naloxone hydrochloride dihydrate | 0054-0189-13 |
| Buprenorphine | 0093-3656-40 |
| Buprenorphine | 0093-3657-40 |
| Buprenorphine | 0093-3658-40 |
| Buprenorphine | 0093-3659-40 |
| Buprenorphine Hydrochloride | 0143-9246-05 |
| Buprenorphine | 0228-3153-03 |
| Buprenorphine and Naloxone | 0228-3154-03 |
| Buprenorphine and Naloxone | 0228-3154-73 |
| Buprenorphine and Naloxone | 0228-3155-03 |
| Buprenorphine and Naloxone | 0228-3155-73 |
| Buprenorphine | 0228-3156-03 |
| Buprenorphine and Naloxone | 0378-8765-93 |
| Buprenorphine and Naloxone | 0378-8766-93 |
| Buprenorphine and Naloxone | 0378-8767-93 |
| Buprenorphine and Naloxone | 0378-8768-93 |
| Buprenorphine HCl and Naloxone HCl | 0406-1923-03 |
| Buprenorphine HCl and Naloxone HCl | 0406-1924-03 |
| Buprenorphine Hydrochloride and Naloxone Hydrochloride Dihydrate | 0406-8005-03 |
| Buprenorphine Hydrochloride and Naloxone Hydrochloride Dihydrate | 0406-8020-03 |
| Buprenorphine Hydrochloride | 0409-2012-32 |
| Buprenorphine Hydrochloride | 0517-0725-05 |
| BUPRENORPHINE AND NALOXONE | 0781-7216-64 |
| BUPRENORPHINE AND NALOXONE | 0781-7227-64 |
| BUPRENORPHINE AND NALOXONE | 0781-7238-64 |
| BUPRENORPHINE AND NALOXONE | 0781-7249-64 |
| buprenorphine hydrochloride and naloxone hydrochloride dihydrate | 0904-7010-06 |
| SUBLOCADE | 12496-0100-1 |
| SUBLOCADE | 12496-0300-1 |
| Buprenex | 12496-0757-5 |
| Suboxone | 12496-1202-3 |
| Suboxone | 12496-1204-3 |
| Suboxone | 12496-1208-3 |
| Suboxone | 12496-1212-3 |
| Buprenorphine Hydrochloride | 42023-179-05 |
| Buprenorphine | 42858-353-40 |
| Buprenorphine | 42858-493-40 |
| BUPRENORPHINE | 42858-501-03 |
| BUPRENORPHINE | 42858-502-03 |
| Buprenorphine | 42858-586-40 |
| Buprenorphine and Naloxone | 42858-601-03 |
| Buprenorphine and Naloxone | 42858-602-03 |
| Buprenorphine | 42858-750-40 |
| Buprenorphine | 42858-839-40 |
| Buprenorphine and Naloxone | 43598-579-30 |
| Buprenorphine and Naloxone | 43598-580-30 |
| Buprenorphine and Naloxone | 43598-581-30 |
| Buprenorphine and Naloxone | 43598-582-30 |
| Buprenorphine and Naloxone | 47781-355-03 |
| Buprenorphine and Naloxone | 47781-356-03 |
| Buprenorphine and Naloxone | 47781-357-03 |
| Buprenorphine and Naloxone | 47781-358-03 |
| Buprenorphine HCl and Naloxone HCl | 50268-144-15 |
| Buprenorphine HCl and Naloxone HCl | 50268-145-15 |
| Buprenorphine and Naloxone | 50383-287-93 |
| Buprenorphine and Naloxone | 50383-294-93 |
| buprenorphine hydrochloride | 50383-924-93 |
| buprenorphine hydrochloride | 50383-930-93 |
| Buprenorphine and Naloxone | 52427-692-03 |
| Buprenorphine and Naloxone | 52427-694-03 |
| Buprenorphine and Naloxone | 52427-698-03 |
| Buprenorphine and Naloxone | 52427-712-03 |
| Probuphine | 52440-100-14 |
| Zubsolv | 54123-114-30 |
| Zubsolv | 54123-907-30 |
| Zubsolv | 54123-914-30 |
| Zubsolv | 54123-929-30 |
| Zubsolv | 54123-957-30 |
| Zubsolv | 54123-986-30 |
| Buprenorphine HCl | 55700-302-30 |
| Buprenorphine | 55700-579-04 |
| BELBUCA | 55700-867-60 |
| Buprenorphine HCl | 58118-0176-8 |
| Buprenorphine HCl | 58118-0177-8 |
| Buprenorphine | 58118-3156-8 |
| Butrans | 59011-750-04 |
| Butrans | 59011-751-04 |
| Butrans | 59011-752-04 |
| Butrans | 59011-757-04 |
| Butrans | 59011-758-04 |
| BUNAVAIL | 59385-012-30 |
| BUNAVAIL | 59385-014-30 |
| BUNAVAIL | 59385-016-30 |
| BELBUCA | 59385-021-60 |
| BELBUCA | 59385-022-60 |
| BELBUCA | 59385-023-60 |
| BELBUCA | 59385-024-60 |
| BELBUCA | 59385-025-60 |
| BELBUCA | 59385-026-60 |
| BELBUCA | 59385-027-60 |
| Buprenorphine and Naloxone | 60429-586-30 |
| Buprenorphine and Naloxone | 60429-587-30 |
| Buprenorphine | 60687-481-21 |
| Buprenorphine | 60687-492-21 |
| Buprenorphine and Naloxone | 62175-452-32 |
| Buprenorphine and Naloxone | 62175-458-32 |
| Buprenorphine | 62756-459-64 |
| Buprenorphine | 62756-459-83 |
| Buprenorphine | 62756-460-64 |
| Buprenorphine | 62756-460-83 |
| Buprenorphine and Naloxone | 62756-969-64 |
| Buprenorphine and Naloxone | 62756-969-83 |
| Buprenorphine and Naloxone | 62756-970-64 |
| Buprenorphine and Naloxone | 62756-970-83 |
| buprenorphine hydrochloride | 63629-7126-1 |
| buprenorphine hydrochloride | 63629-7126-2 |
| buprenorphine hydrochloride | 63629-7126-3 |
| buprenorphine hydrochloride | 63629-7126-4 |
| buprenorphine hydrochloride | 63629-7126-5 |
| buprenorphine hydrochloride | 63629-7126-6 |
| buprenorphine hydrochloride | 63629-7126-7 |
| buprenorphine hydrochloride | 63629-7126-8 |
| buprenorphine hydrochloride | 63629-7126-9 |
| Buprenorphine HCl and Naloxone HCl | 65162-415-03 |
| Buprenorphine HCl and Naloxone HCl | 65162-415-09 |
| Buprenorphine HCl and Naloxone HCl | 65162-416-03 |
| Buprenorphine HCl and Naloxone HCl | 65162-416-09 |
| Buprenorphine | 67046-994-30 |
| Buprenorphine | 67046-995-30 |
| Buprenorphine Hydrochloride Sublingual | 67046-996-30 |
| Buprenorphine Hydrochloride Sublingual | 67046-997-30 |
| Buprenorphine | 69238-1202-2 |
| Buprenorphine | 69238-1203-2 |
| Buprenorphine | 69238-1204-2 |
| Buprenorphine | 69238-1205-2 |
| Buprenorphine | 69238-1505-2 |
| Buprenorphine | 70518-1557-0 |
| Buprenorphine | 70518-1625-0 |
| Buprenorphine HCl and Naloxone HCl | 70518-1684-0 |
| Buprenorphine HCl | 70518-2014-0 |
| buprenorphine hydrochloride | 70518-2216-0 |
| BUPRENORPHINE | 70518-2217-0 |
| BUPRENORPHINE | 70518-2218-0 |
| Buprenorphine HCl | 70518-2226-0 |
| Buprenorphine and Naloxone | 70518-2311-0 |
| Buprenorphine and Naloxone | 70518-2327-0 |
| Buprenorphine | 71335-0353-1 |
| Buprenorphine | 71335-0353-2 |
| Buprenorphine | 71335-0353-3 |
| Buprenorphine | 71335-0353-4 |
| Buprenorphine | 71335-0353-5 |
| Buprenorphine | 71335-0353-6 |
| Buprenorphine | 71335-0353-7 |
| BUPRENORPHINE | 71335-0950-1 |
| BUPRENORPHINE | 71335-0950-2 |
| BUPRENORPHINE | 71335-0950-3 |
| BUPRENORPHINE | 71335-0950-4 |
| BUPRENORPHINE | 71335-0950-5 |
| BUPRENORPHINE | 71335-0950-6 |
| BUPRENORPHINE | 71335-0950-7 |
| BUPRENORPHINE | 71335-1154-1 |
| BUPRENORPHINE | 71335-1154-2 |
| BUPRENORPHINE | 71335-1154-3 |
| BUPRENORPHINE | 71335-1154-4 |
| BUPRENORPHINE | 71335-1154-5 |
| BUPRENORPHINE | 71335-1154-6 |
| BUPRENORPHINE | 71335-1154-7 |
| BUPRENORPHINE | 71335-1154-8 |
| BUPRENORPHINE | 71335-1154-9 |
| Buprenorphine HCl | 71335-1163-1 |
| Buprenorphine HCl | 71335-1163-2 |
| Buprenorphine HCl | 71335-1163-3 |
| Buprenorphine HCl | 71335-1163-4 |
| Buprenorphine HCl | 71335-1163-5 |
| Buprenorphine HCl | 71335-1163-6 |
| Buprenorphine HCl | 71335-1163-7 |
| Buprenorphine HCl | 71335-1163-8 |
| Buprenorphine HCl | 71335-1163-9 |
| Buprenorphine and Naloxone | 71335-1296-1 |
| Buprenorphine and Naloxone | 71335-1296-2 |
| Buprenorphine and Naloxone | 71335-1378-1 |
| Buprenorphine HCl and Naloxone HCl | 71335-1514-1 |
| Buprenorphine HCl and Naloxone HCl | 71335-1514-2 |
| buprenorphine hydrochloride and naloxone hydrochloride dihydrate | 71335-1653-1 |
| buprenorphine hydrochloride and naloxone hydrochloride dihydrate | 71335-1720-1 |
| buprenorphine hydrochloride and naloxone hydrochloride dihydrate | 71335-1720-2 |
| Buprenorphine Hydrochloride and Naloxone Hydrochloride Dihydrate | 71335-1725-1 |

**Supplemental Figure 1.**
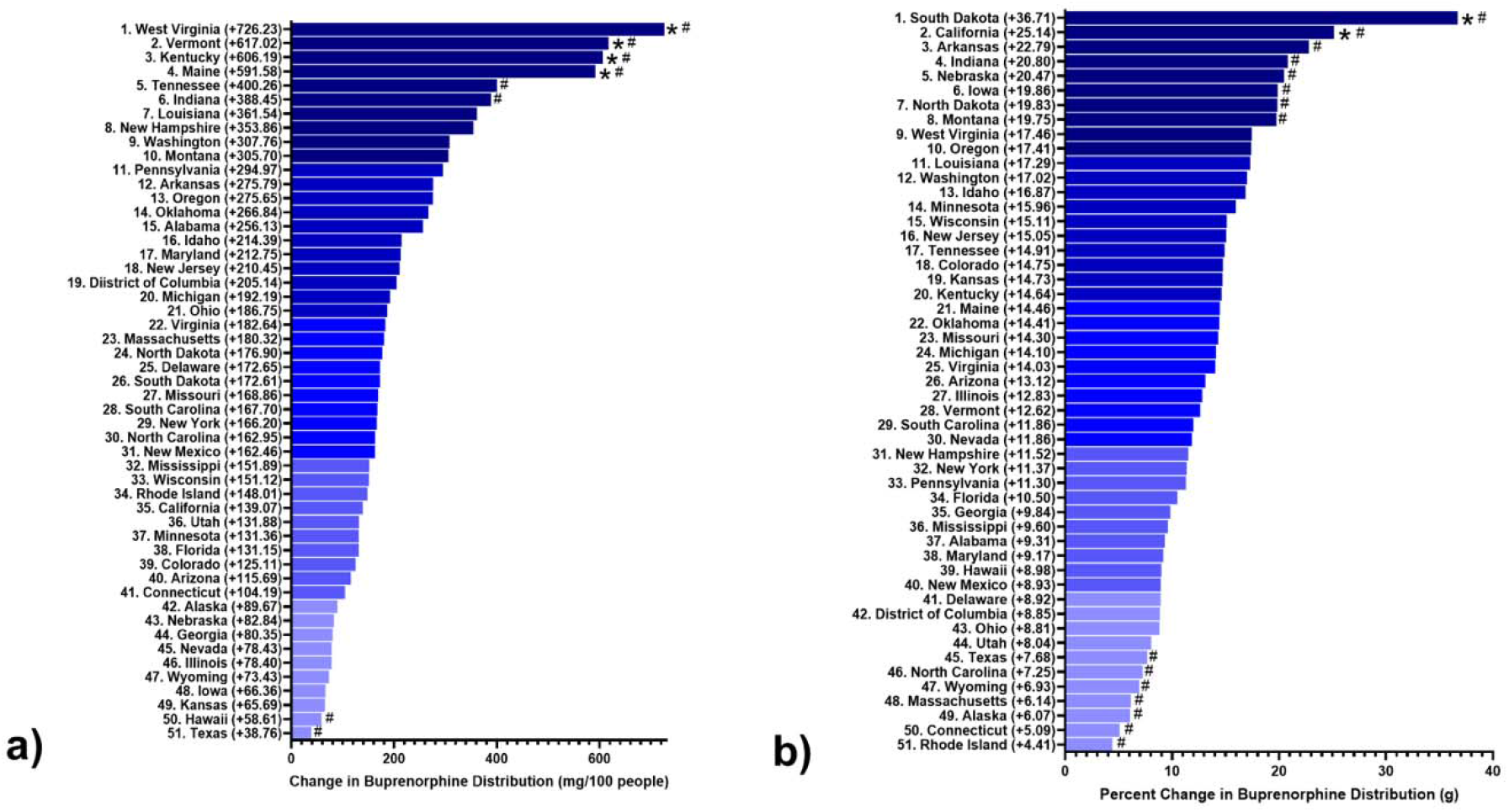
Percentage change buprenorphine distribution by weight per state a) and change in state buprenorphine distribution in mg per 100 people from ARCOS database b) from 2019-20. Percentage change is calculated using the total grams of buprenorphine distributed per state. States that fall outside of a 95% confidence interval from the mean percentage (13.6) and mean change in mg per 100 people (216.0) were noted with an asterisk (*). A pound (#) sign indicates states that were greater than 1 SD from the mean.

**Supplemental Figure 2.**
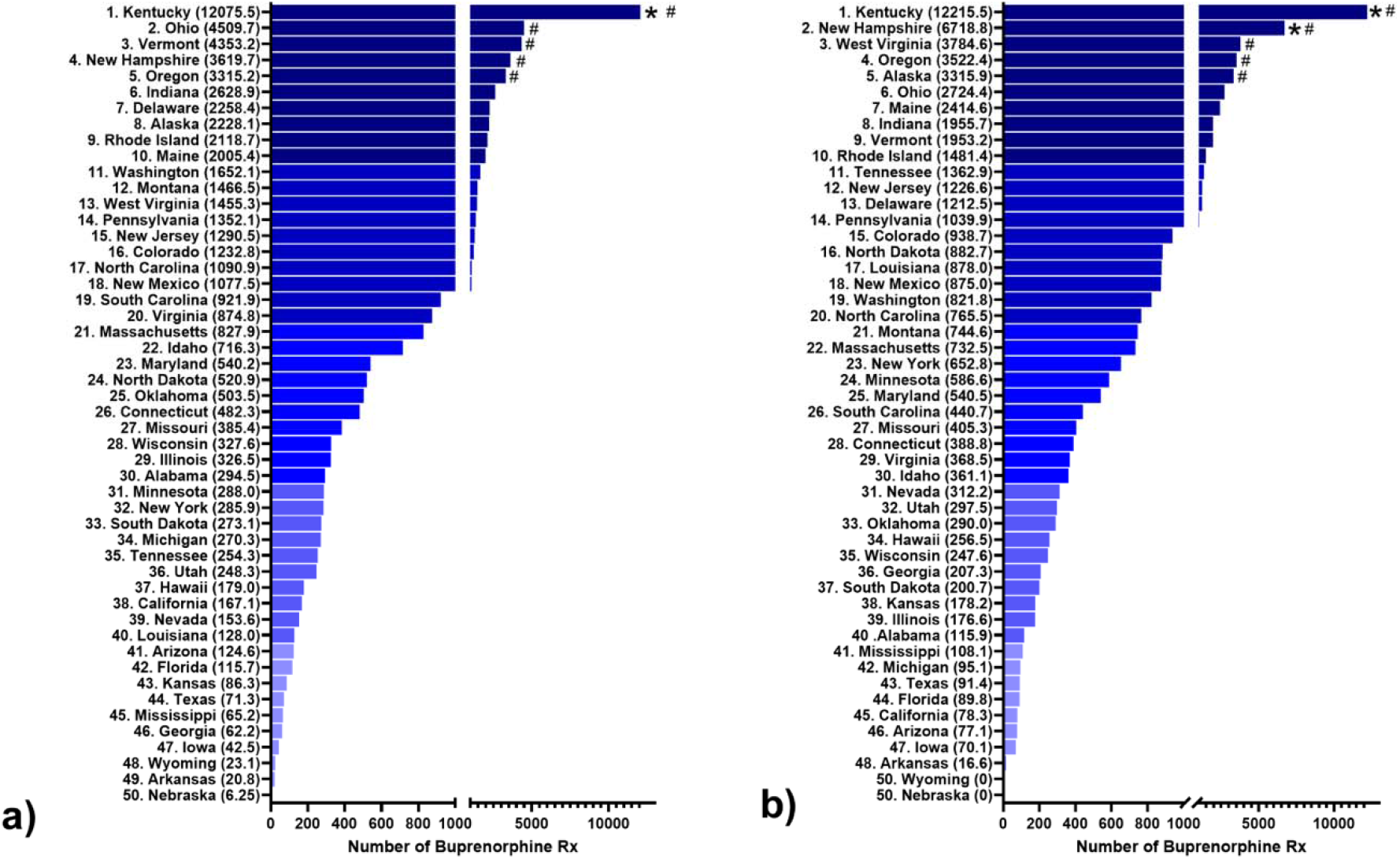
Total number of buprenorphine prescriptions among Medicaid enrollees per state in 2018 a) and 2019 b). Values represent the total number of prescriptions issued normalized to the number of prescriptions per 100,000 Medicaid enrollees in each state. States that fall outside of a 95% CI from the mean number of prescriptions per 100K enrollees (1,186.4 in 2018 and 1,164.4 in 2019) wer annotated with an asterisk (*). States that were greater than 1 SD from the mean were indicated with a pound (#) sign.

**Supplemental Figure 3.**
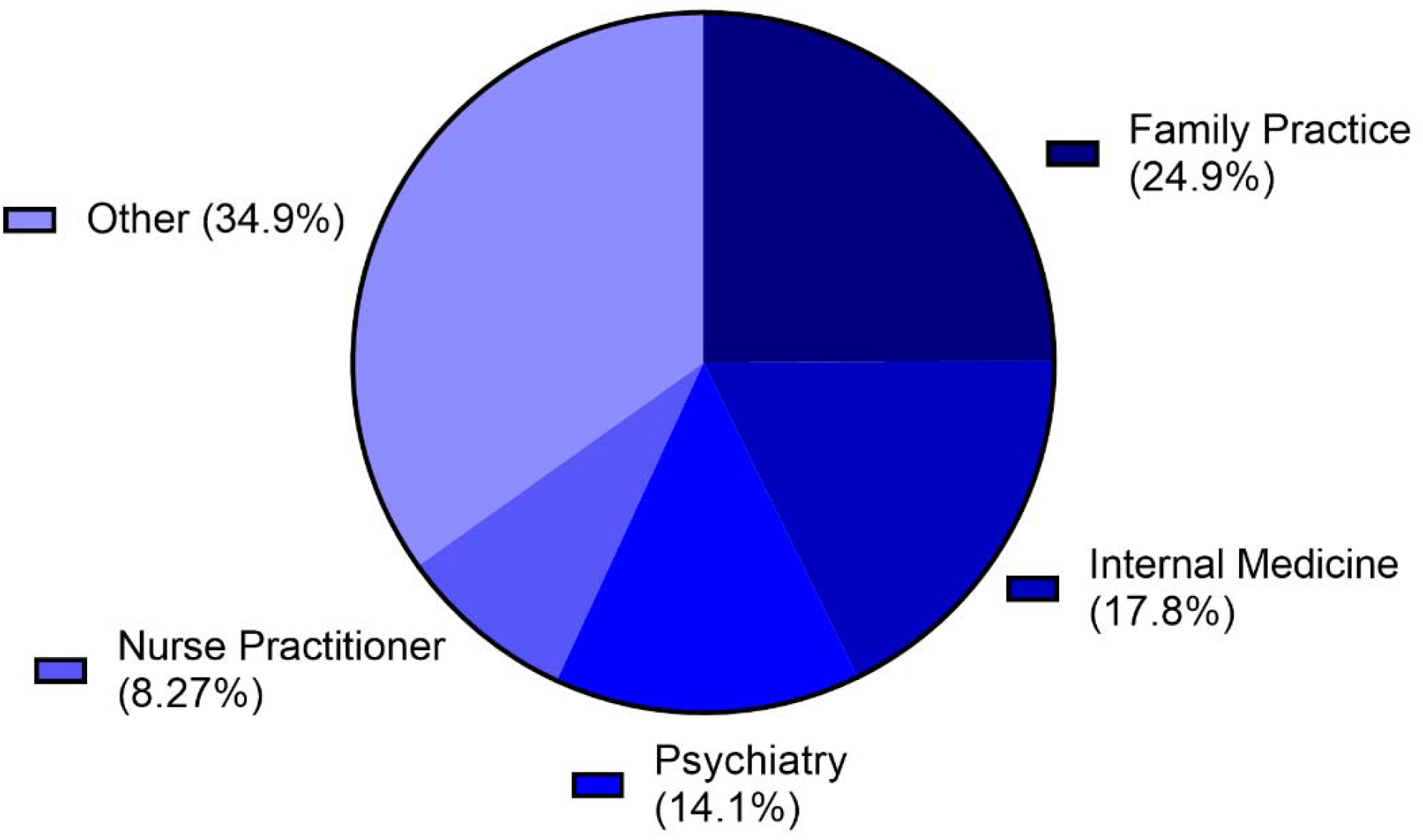
Top prescribers of buprenorphine to Medicare enrollees in 2018 by specialty. Specialty ranking lower than 8.3% were reported as other.

**Supplemental Figure 4.**
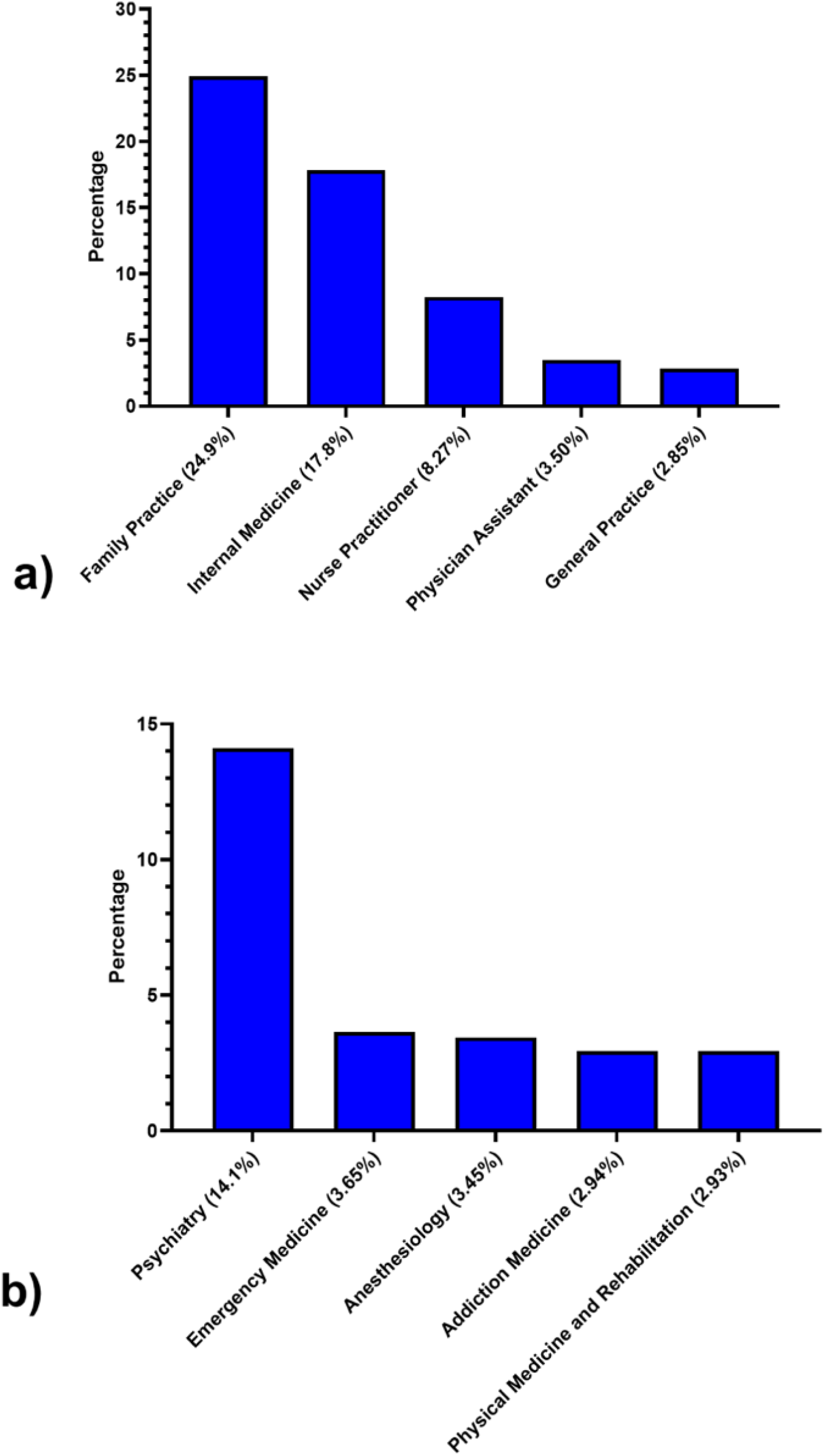
Top prescribers of buprenorphine to Medicare enrollees in 2018 separated to general practitioners a) and specialists b). The specialty categories listed represent exactly how they are reported in the Medicare database.

**Supplemental Figure 5.**
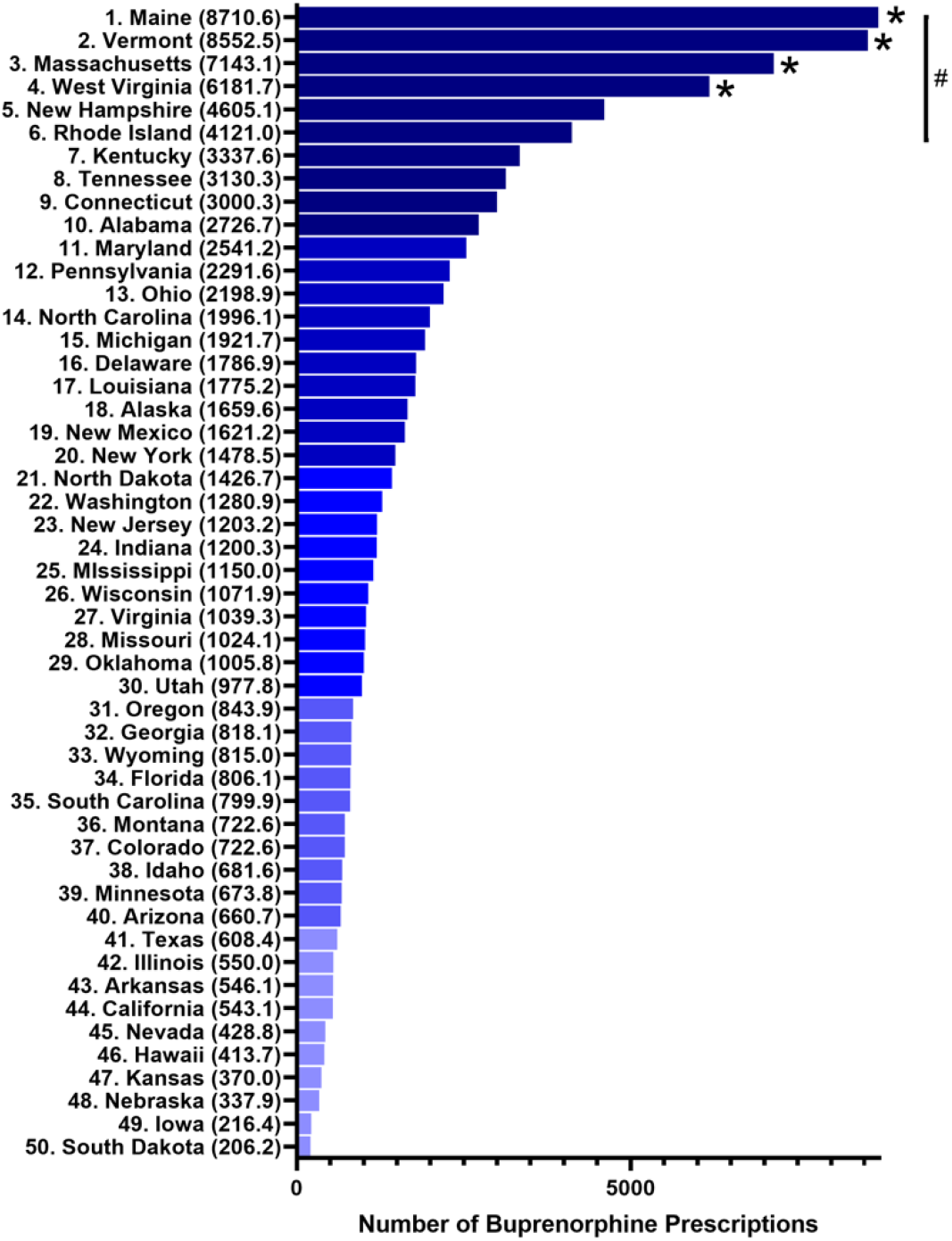
Total number of buprenorphine prescriptions among Medicare enrollees per state in 2018. The values graphed represent the total number of prescriptions issued normalized to the number of prescriptions per 100,000 Medicare enrollees in each state. States that fall outside of a 95% CI from the mean number of prescriptions per 100K enrollees (1,878.5) are annotated with an asterisk (*).

